# Changes in morbimortality patterns due to heart failure in patients of the Unified Health System and their economic impacts: a challenge for Public Health management

**DOI:** 10.1101/2023.03.15.23286880

**Authors:** Matheus Manoel Diogo Lins, Vivian Fernandes Alves Borges

## Abstract

A descriptive ecological study was conducted to form a profile of morbimortality due to cardiac care in the Brazilian population based on patients registered in the Department of Informatics of the Unified Health System (DATASUS). The results of the article show that, during the studied period, 2,984,424 patients were hospitalized due to cardiac care, predominantly represented by males, with the age group predominating in elderly individuals between 70 and 79 years old, of white ethnicity, and with a character of emergency care. The average hospitalization cost was estimated at R$ 1,371.67, generating an annual cost to the Brazilian economy of R$ 341,137,000.

**Summary:** *Background:* Heart failure (HF) is a complex syndrome considered the common endpoint of various cardiovascular diseases, and due to its high prevalence, it is necessary to understand its morbidity and mortality profile.

*Objective:* To describe the morbidity and mortality profile of HF among patients registered in the Department of Informatics of the Unified Health System (DATASUS) and its impact on public health between 2008 and 2020.

*Materials and Methods:* A descriptive epidemiological study was conducted using data obtained from the TABNET application developed by DATASUS. Patients of both sexes, without age restrictions, who were hospitalized with a confirmed diagnosis of HF in Brazil, from January 2008 to December 2020 were included. Data were stratified by sex, age group, self-reported ethnicity/race, type of healthcare plan, and nature of care.

*Results:* During the study period, 2,984,424 patients were hospitalized due to HF in the SUS. The majority were male: 1,533,469 (51.38%); elderly with the predominant age group between 70 and 79 years: 788,367 (26.41%); of white ethnicity: 1,102,860 (36.95%); private healthcare plan: 1,196,559 (46.10%), and with emergency care character: 2,827,354 (94.73%). The average hospitalization cost was estimated at R$ 1,371.67, generating an annual cost to the Brazilian economy of R$ 341,137,000.

*Conclusions:* Heart failure is an important public health problem, causing not only high morbidity in affected individuals but also a significant financial burden on the Unified Health System (SUS).

## INTRODUCTION

Heart failure (HF) is a complex cardiovascular syndrome characterized by the heart’s inability to provide the basic adequate amount of blood to meet metabolic demands, or doing so at the cost of high filling pressures. It is often the end result of various cardiovascular diseases and has a poor prognosis, with mortality rates reaching 50% in the 5 years following diagnosis. It has a multifactorial etiology, with chronic ischemic heart disease associated with systemic arterial hypertension being prominent. However, in regions with low socioeconomic status, it may be related to Chagas disease, endomyocardial fibrosis, and rheumatic valvulopathy. The typical signs and symptoms of HF result from reduced cardiac output and/or high pressures exerted on the cardiac chambers during the filling phase.

HF can be classified based on the following criteria: severity of symptoms (New York Heart Association functional classification - NYHA), ejection fraction (HFpEF, HFmrEF, and HFrEF), and disease progression (various stages). Therapeutic advances observed in the treatment of arterial hypertension, acute myocardial infarction, and HF have allowed for greater survival and improved quality of life for these patients. However, there has been an increase in the incidence of HF, estimated at 1 to 2% of the world’s population, as well as an increase in hospital costs, placing a burden on healthcare systems in various countries, especially those with a growing elderly population. By 2025, the population over 60 years old in Brazil will be the sixth largest in the world, which will reflect in an increase in HF cases and costs for its treatment for the Unified Health System (SUS). However, it is still necessary to deepen these analyses because there is a lack of epidemiological data, which is important for proper planning of preventive methods.

Therefore, recognizing HF as one of the main cardiovascular diseases and its high mortality and costs to public health, it is important to recognize the morbidity profile of HF in Brazil, considering the power of epidemiology in helping to plan effective public policies for the prevention and control of the disease. In order to support this recognition, this article aims to describe the profile of morbidity and mortality due to HF among patients registered in the Department of Health Informatics of the Unified Health System (DATASUS) and its impact on public health between 2008 and 2020.

## MATERIALS AND METHODS

A descriptive ecological epidemiological study was conducted, and the data were obtained through the Department of Health Informatics of the Brazilian Unified Health System (DATASUS), using the Hospital Information System (SIH) platform, accessed in March 2021. patients of both sexes, without age restriction, who were admitted with a confirmed diagnosis of heart failure classified in the category I59.0, Chapter IX-Diseases of the Circulatory System, in the version corresponding to the 10th revision of the International Statistical Classification of Diseases and Related Health Problems (ICD-10), during the period from January 2008 to December 2020, were included. The following variables were analyzed: the number of annual hospitalizations for heart failure, the percentage frequency of elective and emergency admissions, the number of hospitalizations by sex, age group, declared race/color, and type of healthcare (private and public). Statistical analysis The collected data were transcribed and organized into a Microsoft Office Excel (Microsoft©, 2010) spreadsheet for statistical analysis, applying the simple descriptive statistical method, using absolute and relative frequencies for qualitative evaluation. To calculate the mortality rate, the ratio between deaths and hospitalizations for the same disease during the study period was calculated and then multiplied by 100, obtaining the value in percentage.

## Ethical considerations

According to CNS Resolution 510/2016, submission to the Research Ethics Committee was not required due to the nature of the study (ecological).

## RESULTS

Table 1 shows the data on hospitalizations, deaths, and mortality due to heart failure in patients registered in the Department of Health Informatics of the Brazilian Unified Health System (DATASUS) by region and year of service.

**Table 1:** Admissions, deaths, and mortality due to heart failure in patients registered in the Hospital Information System of the Brazilian Unified Health System stratified by region and year of service. Brazil, 2008-2020.

| Variables/<br>year | 2008 | 2009 | 2010 | 2011 | 2012 | 2013 | 2014 | 2015 | 2016 | 2017 | 2018 | 2019 | 2020 | Total | Total |
| --- | --- | --- | --- | --- | --- | --- | --- | --- | --- | --- | --- | --- | --- | --- | --- |
|  | Abs. | Abs. | Abs. | Abs. | Abs. | Abs. | Abs. | Abs. | Abs. | Abs. | Abs. | Abs. | Abs. | Abs. | % |
| Admissions by region |  |  |  |  |  |  |  |  |  |  |  |  |  |  |  |
| North | 15.124 | 14.336 | 13.768 | 13.617 | 13.106 | 12.730 | 11.896 | 11.216 | 10.625 | 10.681 | 11.108 | 11.136 | 8.621 | 157.964 | 5,20% |
| North East | 62.806 | 64.832 | 63.647 | 63.007 | 59.548 | 57.169 | 54.178 | 51.327 | 48.805 | 48.151 | 45.740 | 45.120 | 34.908 | 699.238 | 23,40% |
| Southeast | 112.246 | 114.138 | 110.058 | 111.206 | 103.382 | 98.295 | 93.855 | 91.377 | 89.267 | 86.297 | 81.991 | 82.147 | 72.660 | 1.246.919 | 41,70% |
| South | 57.720 | 58.137 | 57.138 | 54.957 | 50.787 | 50.348 | 47.988 | 48.605 | 49.810 | 48.098 | 47.427 | 47.965 | 41.341 | 660.321 | 22,10% |
| Midwest | 21.091 | 20.493 | 20.191 | 18.915 | 17.590 | 17.901 | 16.460 | 16.378 | 15.953 | 15.497 | 14.567 | 13.476 | 11.470 | 219.982 | 7,30% |
| Total | 268.987 | 271.936 | 264.802 | 261.702 | 244.413 | 236.443 | 224.377 | 218.903 | 214.460 | 208.724 | 200.833 | 199.844 | 169.000 | 2.984.424 | 100% |
|  | Abs. | Abs. | Abs. | Abs. | Abs. | Abs. | Abs. | Abs. | Abs. | Abs. | Abs. | Abs. | Abs. | Abs. | % |
| deaths by region |  |  |  |  |  |  |  |  |  |  |  |  |  |  |  |
| North | 1.021 | 1.014 | 1.145 | 1.201 | 1.192 | 1.117 | 1.061 | 1.124 | 1.320 | 1.205 | 1.250 | 1.430 | 1.126 | 15.206 | 5,13% |
| North East | 4.469 | 4.776 | 4.767 | 5.235 | 5.208 | 5.083 | 5.024 | 5.171 | 5.342 | 5.044 | 5.009 | 5.028 | 4.288 | 64.444 | 21,70% |
| Southeast | 10.979 | 11.579 | 11.856 | 12.086 | 11.200 | 10.796 | 10.307 | 10.609 | 10.951 | 10.783 | 10.332 | 10.561 | 9.905 | 141.944 | 47,90% |
| South | 4.050 | 4.200 | 4.344 | 4.331 | 4.002 | 4.370 | 4.103 | 4.172 | 4.367 | 4.138 | 4.345 | 4.336 | 4.009 | 54.767 | 18,40% |
| Midwest | 1.561 | 1.600 | 1.640 | 1.533 | 1.517 | 1.582 | 1.557 | 1.697 | 1.635 | 1.486 | 1.401 | 1.451 | 1.145 | 19.805 | 6,60% |
| Total | 22.080 | 23.169 | 23.752 | 24.386 | 23.119 | 22.948 | 22.052 | 22.773 | 23.615 | 22.656 | 22.337 | 22.806 | 20.473 | 296.166 | 100% |
| Mortality rate |  |  |  |  |  |  |  |  |  |  |  |  |  |  |  |
| North | 8,21 | 8,52 | 8,97 | 9,32 | 9,46 | 9,71 | 9,83 | 10,40 | 11,01 | 10,85 | 11,12 | 11,12 | 11,41 | 9,92 | * |
| North East | 6,75 | 7,07 | 8,32 | 8,82 | 9,10 | 8,77 | 8,92 | 10,02 | 12,42 | 11,28 | 11,25 | 12,84 | 13,06 | 9,63 | * |
| Southeast | 9,78 | 10,14 | 10,77 | 10,87 | 10,83 | 10,98 | 10,98 | 11,61 | 12,27 | 12,50 | 12,60 | 12,86 | 13,63 | 11,38 | * |
| South | 7,02 | 7,22 | 7,60 | 7,88 | 7,88 | 8,68 | 8,55 | 8,58 | 8,77 | 8,60 | 9,16 | 9,04 | 9,70 | 8,29 | * |
| Midwest | 7,40 | 7,81 | 8,12 | 8,10 | 8,62 | 8,84 | 9,46 | 10,36 | 10,25 | 9,59 | 9,62 | 10,77 | 9,98 | 9,00 | * |
| Total | 8,21 | 8,52 | 8,97 | 9,32 | 9,46 | 9,71 | 9,83 | 10,40 | 11,01 | 10,85 | 11,12 | 11,41 | 12,11 | 9,92 | * |

During the studied period, there were 2,984,424 hospitalizations and 296,166 deaths, corresponding to a mortality rate of 9.92%.

The southeastern region of the country showed a higher percentage of hospitalizations (41.70%), deaths (47.90%), and mortality rate (11.38%).

The total number of hospitalizations for heart failure among patients registered in DATASUS decreased by approximately 62.82% between 2008 and 2020. Meanwhile, the mortality rate increased by about 38.97% in the same period for the same patients.

Regarding age groups, the population aged between 70 and 79 years old accounted for the highest percentage of hospitalizations (26.41%). As for deaths and mortality rates, the age group ≥ 80 years old was predominant, with 32.41% and 14.99%, respectively, according to Table 2.

**Table 2:** Admissions, deaths, and mortality due to heart failure in patients registered in the Brazilian Unified Health System Hospital Information System stratified by age group. Brazil, 2008 to 2020 (absolute and relative frequencies).

| Age group | Admissions |  | Deaths |  | Mortality rate |
| --- | --- | --- | --- | --- | --- |
|  | Abs. | % | Abs. | % |  |
| <1 ano | 17.565 | 0,58 | 1.902 | 0,64 | 10,83 |
| 1 a 4 anos | 9.997 | 0,33 | 468 | 0,15 | 4,68 |
| 5 a 9 anos | 5.921 | 0,19 | 245 | 0,08 | 4,14 |
| 10 a 14 anos | 6.966 | 0,23 | 423 | 0,14 | 6,07 |
| 15 a 19 anos | 10.473 | 0,35 | 804 | 0,27 | 7,68 |
| 20 a 29 anos | 35.616 | 1,19 | 2.687 | 0,90 | 7,54 |
| 30 a 39 anos | 85.022 | 2,84 | 5.872 | 1,98 | 6,91 |
| 40 a 49 anos | 212.978 | 7,13 | 13.887 | 4,68 | 6,52 |
| 50 a 59 anos | 468.622 | 15,70 | 33.013 | 11,14 | 7,04 |
| 60 a 69 anos | 702.398 | 23,53 | 59.164 | 19,97 | 8,42 |
| 70 a 79 anos | 788.367 | 26,41 | 81.692 | 27,58 | 10,36 |
| $\geq 80$ anos | 640.499 | 21,46 | 96.009 | 32,41 | 14,99 |
| Total | 2.984.424 | 100 | 296.166 | 100 | 9,92 |

In relation to gender, Table 3 shows that the majority of hospitalizations occur in the male population. However, the number of deaths and the mortality rate were higher in the female gender.

**Table 3:**
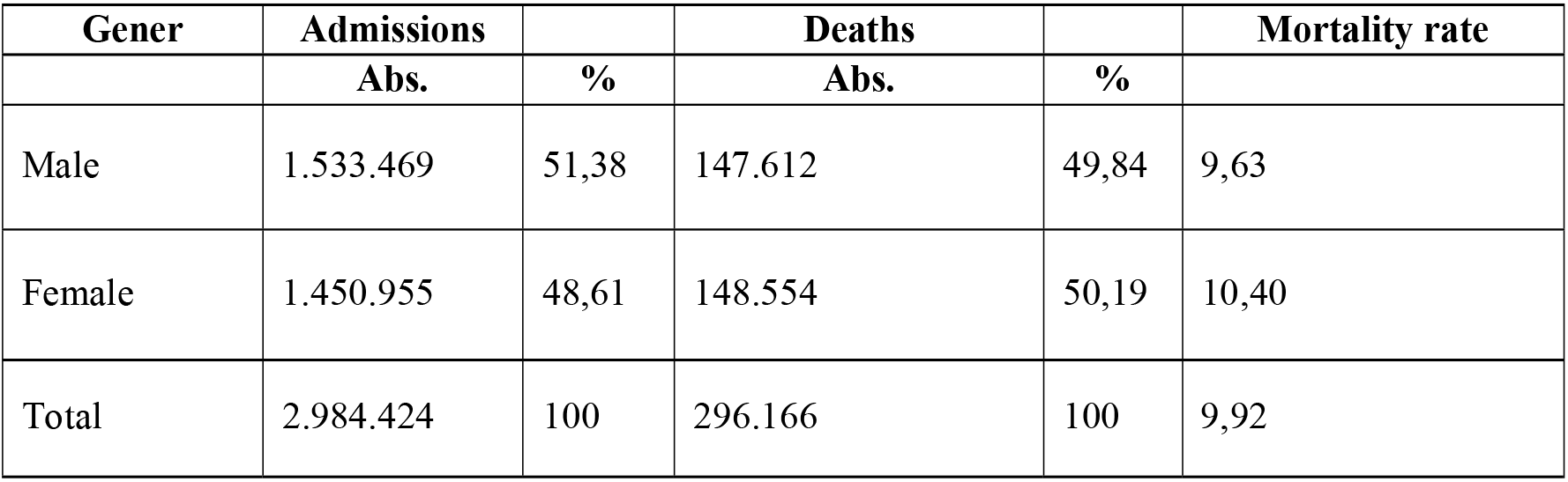
Admissions, deaths and mortality due to heart failure in patients registered in the Hospital Information System of the Brazilian Unified Health System stratified by gender. Brazil, 2008-2020.

| Gener | Admissions |  | Deaths |  | Mortality rate |
| --- | --- | --- | --- | --- | --- |
|  | Abs. | % | Abs. | % |  |
| Male | 1.533.469 | 51,38 | 147.612 | 49,84 | 9,63 |
| Female | 1.450.955 | 48,61 | 148.554 | 50,19 | 10,40 |
| Total | 2.984.424 | 100 | 296.166 | 100 | 9,92 |

In relation to race/ethnicity by year, Table 4 shows that white individuals had the highest rates of hospitalizations and deaths during the analyzed period. However, in terms of mortality rate, the “unspecified” group had the highest numbers, followed by the yellow race. Indigenous people had the lowest mortality rate.

**Table 4:** Admissions, deaths, and mortality due to heart failure in patients registered in the Brazilian Unified Health System (SUS) stratified by race/ethnicity. Brazil, 2008-2020.

| Race/ethnicity | Admissions |  | Deaths |  | Mortality rate |
| --- | --- | --- | --- | --- | --- |

|  | **Abs.** | **%** | **Abs.** | **%** |  |
| --- | --- | --- | --- | --- | --- |
| White | 1.102.860 | 36,95 | 109.900 | 37,10 | 9,97 |
| Black | 137.410 | 4,60 | 12.963 | 4,37 | 9,43 |
| Yellow race | 856.198 | 28,68 | 77.770 | 26,25 | 9,08 |
| Amarela | 28.255 | 0,94 | 2.881 | 0,97 | 10,20 |
| indigenous | 3.543 | 0,11 | 279 | 0,09 | 7,87 |
| no information | 856.158 | 28,68 | 92.373 | 31,18 | 10,79 |
| Total | 2.984.424 | 100 | 296.166 | 100 | 9,92 |

The table 5 shows that the public healthcare system presented the highest mortality rate (11.33%). Regarding the number of hospitalizations, the majority occurred in the private healthcare system (40.09%). The group with no information on the type of hospitalization, described as “Unknown” had the highest number of deaths (38.99%).

**Table 5:** Admissions, deaths, and mortality rates due to heart failure in patients registered in the Brazilian Public Healthcare System stratified by healthcare system type. Brazil, 2008-2020.

| **Regime** | **Admissions** |  | **Deaths** |  | **Mortality rate** |
| --- | --- | --- | --- | --- | --- |
|  | **Abs** | **%** | **Abs** | **%** | **Abs** |
| Public | 759.818 | 25,45 | 86.111 | 29,07 | 11,33 |
| Private | 1.196.559 | 40,09 | 94.565 | 31,92 | 7,90 |
| Unknown | 1.028.047 | 34,44 | 115.490 | 38,99 | 11,23 |
| Total | 2.984.429 | 100 | 296.166 | 100 | 9,92 |

Table 6 shows that cases attended to emergencies had the highest mortality rate (10.02%), as well as the highest number of hospitalizations (94.74%) and deaths (95.65%).

**Table 6:** Hospitalizations, deaths, and mortality rates for heart failure patients registered in the Hospital Information System of the Brazilian Unified Health System, stratified by type of care. Brazil, 2008-2020.

| **Category** | **Admissions** |  | **Deaths** |  | **Mortality rate** |
| --- | --- | --- | --- | --- | --- |
|  | **Abs** | **%** | **Abs** | **%** | **Abs** |
| Elective | 156.520 | 5,24 | 12.815 | 4,32 | 8,19 |
| Urgency | 2.827.354 | 94,74 | 283.297 | 95,65 | 10,02 |
| Others | 550 | 0,01 | 54 | 0,01 | 9,81 |
| Total | 2.984.424 | 100 | 296.166 | 100 | 9,92 |

Table 7 shows an increase in the time (↑5.38%) and average value of hospitalization (↑105%) between 2008 and 2020. The mortality rate also increased during this period (↑47.50%).

**Table 7:**
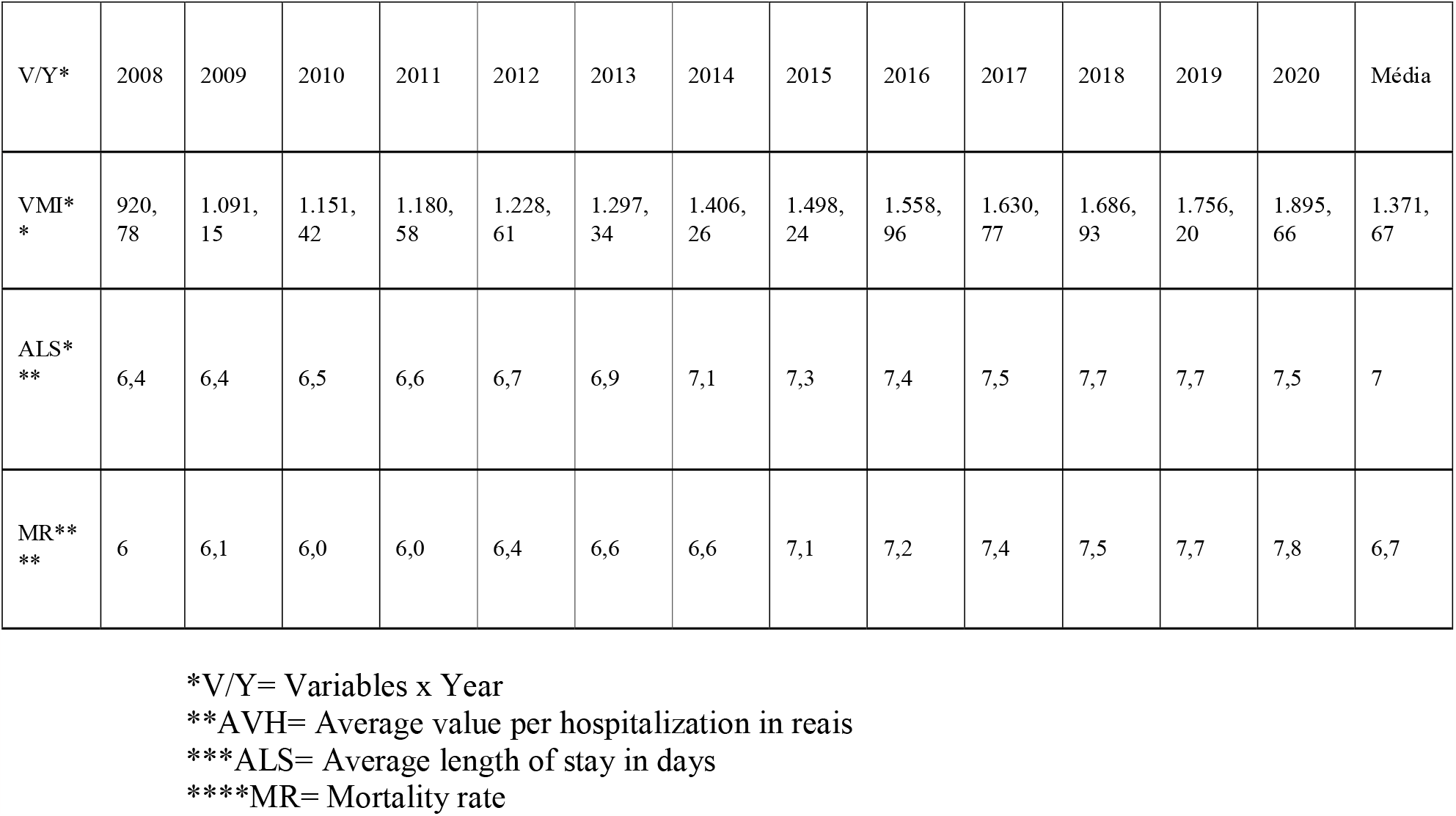
Average value per admission and average length of stay for heart failure patients registered in the Hospital Information System of the Brazilian Unified Health System (SUS), 2008-2020.

## DISCUSSION

Morbidity refers to the number of individuals in a population affected by a particular disease or health problem. The morbidity patterns of a population can be measured through hospital admissions, which are thus called hospital morbidity8.

Morbidity is a widely used parameter for epidemiological surveillance, organization, and public health service interventions9.

Population aging is already a reality in Brazilian society and also a risk factor for the development of chronic diseases due to biological changes, especially in the circulatory system. Studies suggest that by 2025, the population over 60 years old in Brazil will be the sixth largest in the world7, and it is expected that approximately 2% of the Brazilian population will develop IC with an incidence of 200 thousand new cases per year3,6. The demographic transition leads to the epidemiological transition, meaning the exchange of the population’s disease profile, forcing us to learn how to control elderly people’s diseases. This population aging will likely be reflected in the increase in cases of IC and in the increase in costs to the SUS. It was observed that IC presented, during the studied period, an average value per hospitalization of R$ 1,371.67, generating an average annual economic impact of R$ 341,137,000. The increase in the cost of hospitalization of patients with IC was from R$920.78 in 2008 to R$1,895.66 in 2020, an increase of about 105%. Although there is a tendency to think that greater resources lead to better results, this was not observed when analyzing the data from table 7, which shows an increase in the average length of hospital stay and the mortality rate during the same period. This cost increase may be related to several causes, including greater severity of patients, justifying the increase in the average length of hospital stay and the number of deaths observed. The multifactorial cause of the increase in the average value includes, in addition to the increase in the average length of hospital stay, the implementation of new therapeutic methods such as the use of stents and cardiac resynchronization, the increase in the performance of cardiac surgeries, and the introduction of new drugs such as angiotensin-converting enzyme inhibitors (ACEIs) and angiotensin receptor blockers (ARBs) 3,10-12.

In addition, the increase in IPCA, one of the main inflation indicators, during this period (2008-2020) was 103.56%, close to the cost increase observed by the study.

High mortality rates in the age group under 1 year occur mainly due to congenital heart diseases, which are the second leading cause of death in children under 1 year, with an estimated incidence of 1 new case for every 100 births, totaling approximately 28,900 children per year, where up to 80% require cardiac surgery30.

The increase in the in-hospital mortality rate observed in the study may be a result of the paradigm shift in the disease, with the introduction of new therapies such as the use of beta-blockers and spironolactone, drugs with potential for reducing sudden death (a cause of death in up to a third of patients with IC 3,14,15). These drugs increase the survival of severely ill patients. However, as age, pathology, and heart failure progress, these patients tend to die at a later age and in hospital, transferring these deaths, previously extrahospital, to the intra-hospital ones10,11.

It is known that the better the adherence to treatment, the less the need for emergency cardiovascular care16 and that the need for the use of more than two drugs increases the chances of non-adherence by 2.59%.

## CONCLUSIONS

Thus, the study demonstrated that the morbidity profile of heart failure patients registered in the Hospital Information System of the Unified Health System (SUS) is of men, aged 70 to 79, of white ethnicity, from the southeast region, treated under private regime and on an urgent basis. Heart failure is an important public health problem, causing, in addition to a high financial cost to the Unified Health System (SUS), a high morbidity in affected individuals. The study contributes to expanding the analysis of the epidemiological scenario of heart failure patients assisted by SUS, bringing important variables within the context of Brazilian public health, generating evidence and enabling better quality of healthcare provision by health professionals as well as support for the debate of new public policies by authorities and the population aimed at prevention and universal access to health.

## Data Availability

All data produced in the present work are contained in the manuscript

